# Stability of the COVID-19 virus under wet, dry and acidic conditions

**DOI:** 10.1101/2020.04.09.20058875

**Authors:** Zhi-ping Sun, Xia Cai, Chen-jian Gu, Rong Zhang, Wen-dong Han, Yun Qian, Yu-yan Wang, Wei Xu, Yang Wu, Xunjia Cheng, Zheng-hong Yuan, You-hua Xie, Di Qu

## Abstract

COVID-19 has become a pandemic and is spreading fast worldwide. The COVID-19 virus is transmitted mainly through respiratory droplets and close contact. However, the fecal-oral transmission of the virus has not been ruled out and it is important to ascertain how acidic condition in the stomach affects the infectivity of the virus. Besides, it is unclear how stable the COVID-19 virus is under dry and wet conditions. In the present study, we have shown that the COVID-19 virus is extremely infectious as manifested by the infection of Vero-E6 cells by one PFU (Plaque Forming Unit) of the virus. We then investigated the stability of the COVID-19 virus in wet, dry and acidic (pH2.2) environments at room temperature. Results showed that the COVID-19 virus could survive for three days in wet and dry environments, but the dry condition is less favorable for the survival of the virus. Our study also demonstrated that the COVID-19 virus at a relative high titer (1.2 x 103 PFU) exhibits a certain degree of tolerance to acidic environment at least for 60 minutes. When the virus titer was ≤1.0 x 103 PFU, acid treatment (pH2.2) for 30 or 60 minute resulted in virus inactivation. It suggests that the virus at a high concentration may survive in the acidic environment of the stomach. The finding of the present study will contribute to the control of the spread of the COVID-19 virus.

---

To the Editor,

An outbreak of pneumonia caused by a novel coronavirus was reported in December 2019 in Wuhan, Hubei Province of China ^[1-3]^. The COVID-19 virus (SARS-COV-2) has spread to every continent and WHO declared COVID-19 as a pandemic ^[4]^. In this study, the presence of CPE was used as the main index for the detection of the infectivity of the COVID-19 virus. When CPE was not observed, immunofluorescence of intracellular viral N protein and RT-PCR of viral RNA in the cell culture supernatant were used for the detection of viral infection. The strain nCoV-SH01 (GenBank accession no. MT121215) used in this study was isolated and plaque -purified from the nasal-pharyngeal swab of a clinically confirmed COVID-19 patient in Shanghai ^[5]^.

Using the strain nCoV-SH01 and these criteria, we first investigated the infectivity of the COVID-19 virus. We found that as the virus titer decreases (2000, 1000, 500, 250, 100, 50, 10, 5, and 1 PFU), the time for CPE to appear is delayed. One PFU is able to cause infection of Vero-E6 cells, resulting in obvious CPE at 72 hours, whereas higher titer of the virus induced CPE at 24 hours post inoculation (Table 1). It indicates that the COVID-19 virus is highly infectious, and underscores the challenge to control the spread of the COVID-19 virus.

**Table 1.**
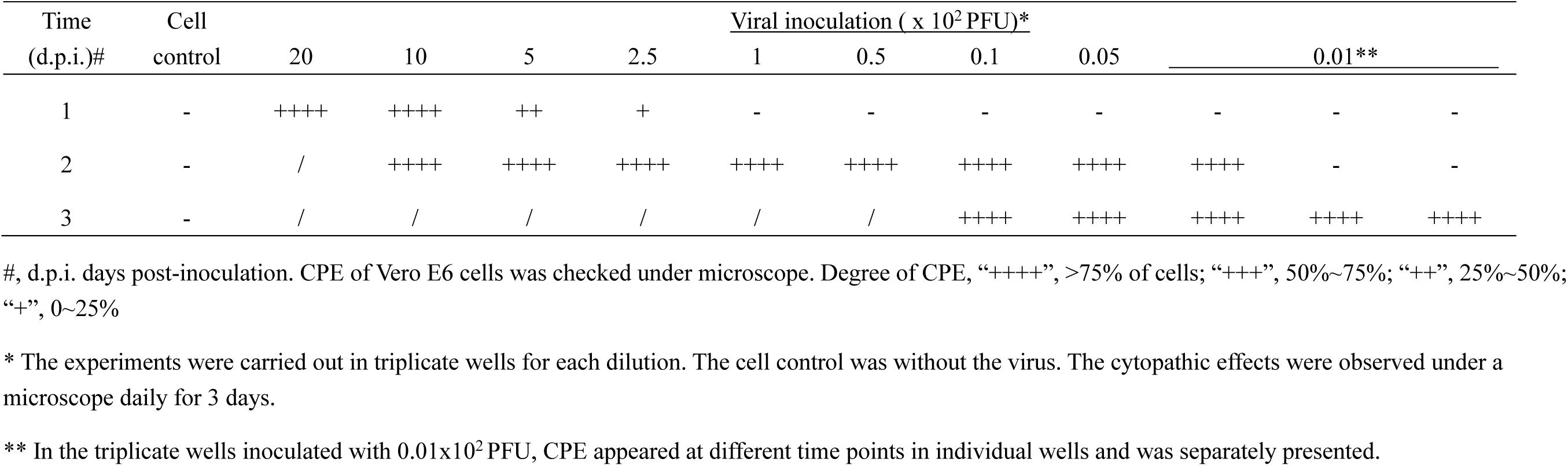
The cytopathic effect of Vero-E6 cell infected by different PFU of the COVID-19 virus

We then studied the stability of the COVID-19 virus in the wet (in 100 uL culture medium) and dry (10 uL supernatant on filter paper) environments at room temperature (22°C) for 1, 2, 3, 4, 5, 6, 7 days respectively. Our results show that the COVID-19 virus can survive for 3 days in the wet or dry environment investigated in this study. Although the virus maintained its infectivity within 3 days in the dry condition, CPE appeared later than that kept in the wet environment, indicating that the dry environment may be less favorable for the survival of the COVID-19 virus (Table 2 & Table 3). However, when the virus had been kept in the wet or dry condition for more than 4 days, no CPE was observed (Table 2 & Table 3), which was confirmed by immune florescence staining (data not shown) with the antibody against viral N protein as well as qRT-PCR.

**Table 2.**
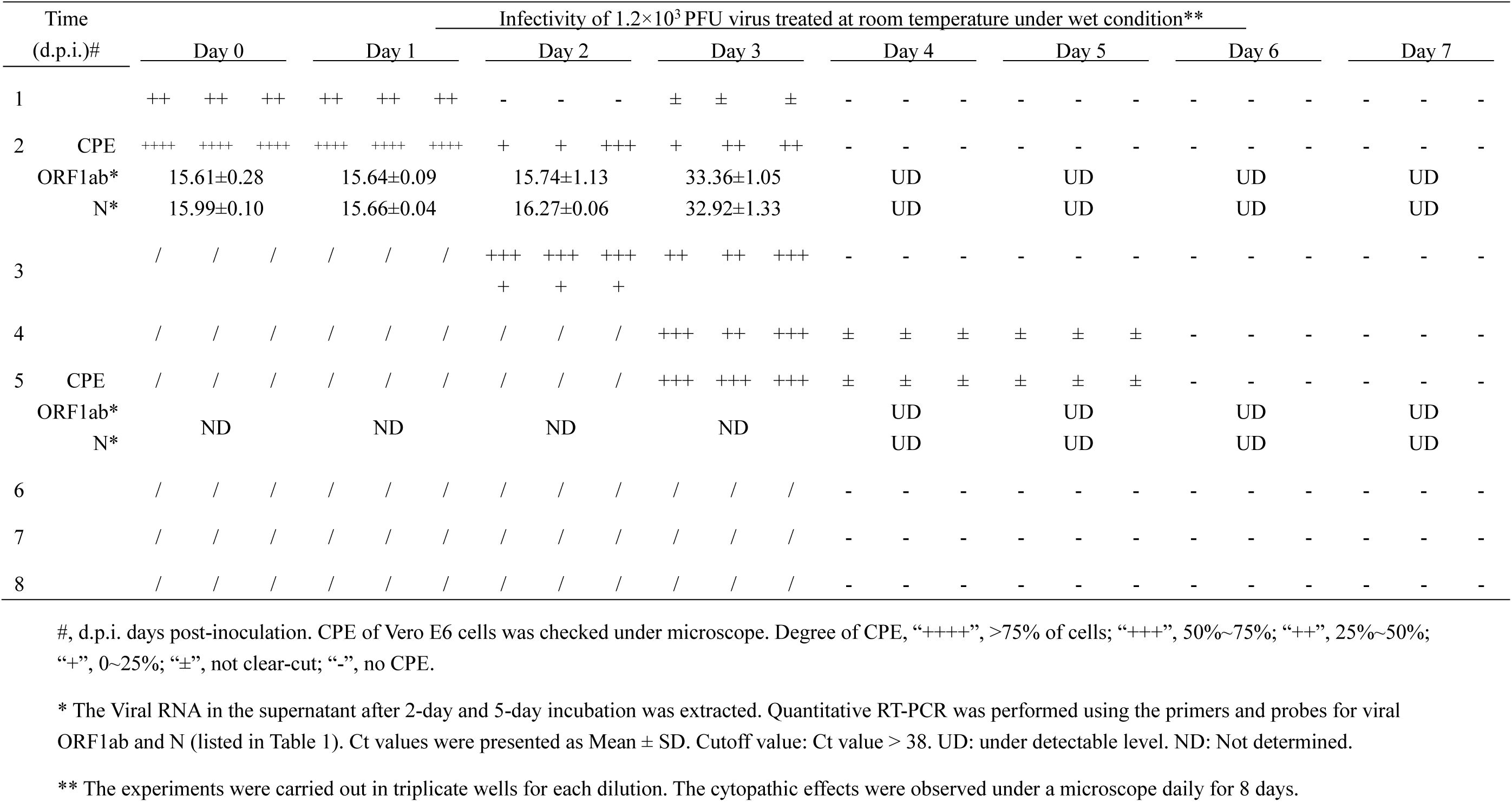
Stability of the COVID-19 virus under wet condition

**Table 3.** Stability of the COVID-19 virus under dry condition.

| Time<br>(d.p.i.)# |  | Infectivity of 1.2×10<sup>3</sup> PFU virus treated at room temperature under dry condition** |  |  |  |  |  |  |  |  |  |  |  |  |  |  |  |  |  |  |  |  |  |  |  |
| --- | --- | --- | --- | --- | --- | --- | --- | --- | --- | --- | --- | --- | --- | --- | --- | --- | --- | --- | --- | --- | --- | --- | --- | --- | --- |
|  |  | Day 0 |  |  | Day 1 |  |  | Day 2 |  |  | Day 3 |  |  | Day 4 |  |  | Day 5 |  |  | Day 6 |  |  | Day 7 |  |  |
| 1 |  | - | - | - | ± | ± | ± | - | - | - | - | - | - | - | - | - | - | - | - | - | - | - | - | - | - |
| 2 | CPE | ++++ | ++++ | ++++ | + | + | + | + | - | + | ± | ± | ± | - | - | - | - | - | - | - | - | - | - | - | - |
|  | ORF1ab* | 15.25±0.22 |  |  | 29.05±0.45 |  |  | 35.09±0.18 |  |  | 35.74±0.09 |  |  | UD |  |  | UD |  |  | UD |  |  | UD |  |  |
|  | N* | 15.06±0.59 |  |  | 28.60±0.15 |  |  | 35.12±0.42 |  |  | 35.98±0.19 |  |  | UD |  |  | UD |  |  | UD |  |  | UD |  |  |
| 3 |  | / | / | / | + | + | + | + | + | + | ± | ± | ± | - | - | - | - | - | - | - | - | - | - | - | - |
| 4 |  | / | / | / | ++++ | ++ | + | + | + | + | ± | ± | ± | ± | ± | - | - | - | - | - | - | - | - | - | - |
| 5 | CPE | / | / | / | ++++ | ++ | ++ | ++ | ++ | + | + | + | + | ± | ± | ± | ± | ± | ± | - | - | - | - | - | - |
|  | ORF1ab* | ND |  |  | ND |  |  | ND |  |  | ND |  |  | UD |  |  | UD |  |  | UD |  |  | UD |  |  |
|  | N* | ND |  |  | ND |  |  | ND |  |  | ND |  |  | UD |  |  | UD |  |  | UD |  |  | UD |  |  |
| 6 |  | / | / | / | / | +++ | +++ | +++ | +++ | + | ++ | ++ | ++ | - | - | - | - | - | - | - | - | - | - | - | - |
| 7 |  | / | / | / | / | +++ | +++ | +++ | +++ | + | ++ | ++ | ++ | - | - | - | - | - | - | - | - | - | - | - | - |
| 8 |  | / | / | / | / | / | / | / | / | + | ++ | ++ | ++ | - | - | - | - | - | - | - | - | - | - | - | - |

#, d.p.i. days post-inoculation. CPE of Vero E6 cells was checked under microscope. Degree of CPE, “++++”, >75% of cells; “+++”, 50%~75%; “++”, 25%~50%; “+”, 0~25%; “±”, not clear-cut; “-”, no CPE.
\* The Viral RNA in the supernatant after 2-day and 5-day incubation was extracted. Quantitative RT-PCR was performed using the primers and probes for viral ORF1ab and N (listed in Table 1). Ct values were presented as Mean ± SD. Cutoff value: Ct value > 38. UD: under detectable level. ND: Not determined.
\*\* The experiments were carried out in triplicate wells for each dilution. The cytopathic effects were observed under a microscope daily for 8 days.

We further investigated the stability of the COVID-19 virus at pH2.2 condition. It shows that the COVID-19 virus has a certain degree of tolerance to acidic environment. In the present study, when 1.2 × 10^3^ PFU of the COVID-19 virus were treated with acidic saline of pH2.2 for 30 or 60 minutes, it still resulted in CPE in the cells, whereas 1.0 × 10^3^ PFU of the COVID-19 virus treated with pH2.2 saline for 30 or 60 minute, no CPE were observed (Table 4). It suggests under the acidic condition the COVID-19 virus at a relatively high titer can survive under acidic condition for at least 1 hour.

**Table 4.**
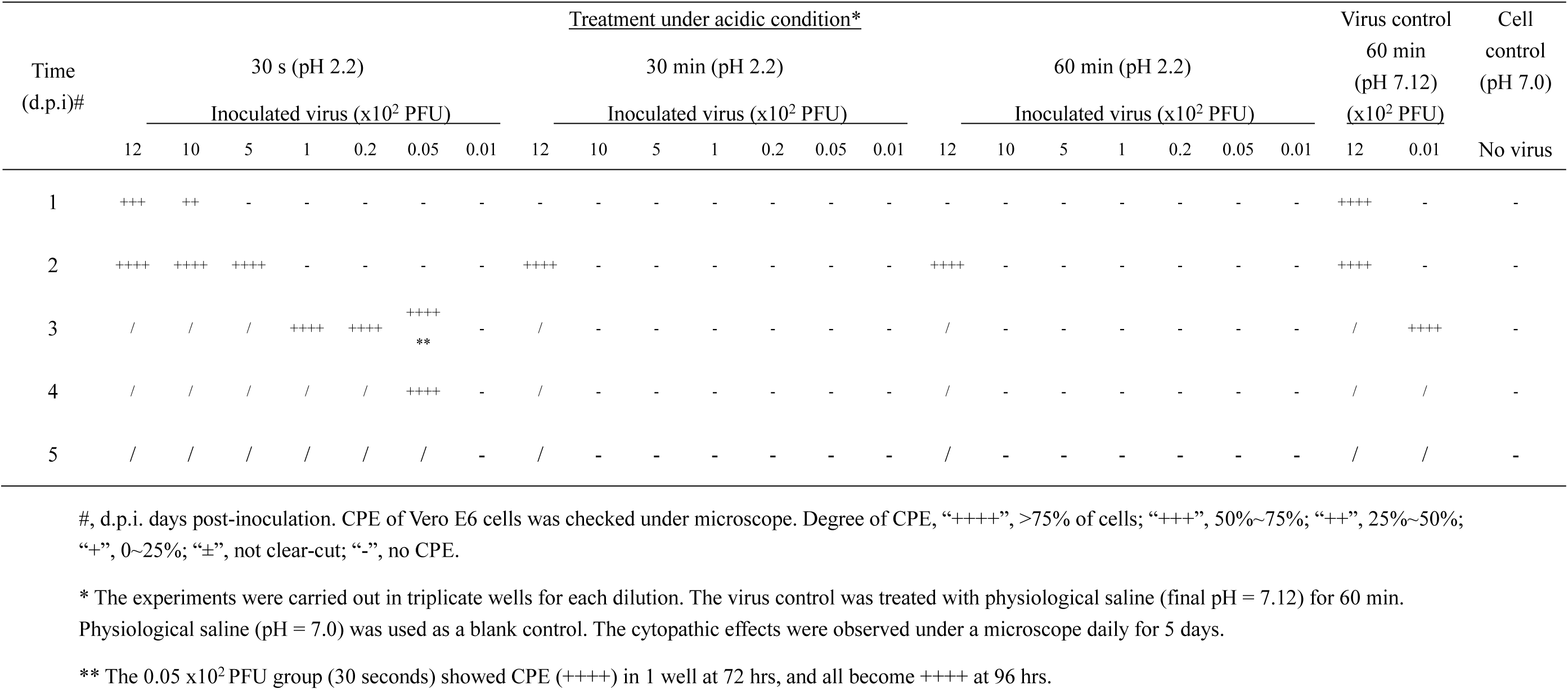
Stability of the COVID-19 virus under acidic condition

In conclusion, our findings show that the COVID-19 virus is highly infectious that one PFU can results in cell infection in our *in vitro* system, and can survive for 3 days in the wet or dry environment. In addition, we for the first time demonstrated that the COVID-19 virus at relative high concentrations can survive under acidic condition that minic the gastric environment. The study would also provide guidance on taking appropriate measures to control the spread of the COVID-19 virus and improve laboratory safety.

The study was supported by the National Science and Technology Major Project (NSTMP) for the Prevention and Treatment of Infectious Diseases (2018ZX10734401, 2018ZX10301208), NSTMP for the Development of Novel Drugs (2019ZX09721001), and Project of Novel Coronavirus Research of Fudan University.

## Data Availability

The data used to support the findings of this study are included within the article

## Reference

1. Wang, C., et al., A novel coronavirus outbreak of global health concern. Lancet, 2020. 395(10223): p. 470–473.

2. Qun Li, X.G., Peng Wu, Xiaoye Wang, Lei Zhou, Yeqing Tong, Ruiqi Ren, Kathy S.M. Leung, EricH.Y. Lau, Jessica Y. Wong, Xuesen Xing, Nijuan Xiang, Yang Wu, Chao Li, Qi Chen, Dan Li, Tian Liu, Jing Zhao, Man Liu, Wenxiao Tu, Chuding Chen, Lianmei Jin, Rui Yang, Qi Wang, Suhua Zhou, Rui Wang, Hui Liu, Yinbo Luo, Yuan Liu, Ge Shao, Huan Li, Zhongfa Tao, Yang Yang, Zhiqiang Deng, Boxi Liu, Zhitao Ma, Yanping Zhang, Guoqing Shi, Tommy T.Y. Lam, Joseph T. Wu, George F. Gao, D. Phil., Benjamin J. Cowling, Bo Yang, Gabriel M. Leung, Zijian Feng, Early Transmission Dynamics in Wuhan, China, of Novel Coronavirus–Infected Pneumonia. The New England Journal of Medicine, 2020(382): p. 1199.

3. The 2019-nCoV Outbreak Joint Field Epidemiology Investigation Team, Q.L., Notes from the Field: An Outbreak of NCIP (2019-nCoV) Infection in China — Wuhan, Hubei Province, 2019- 2020. China CDC Weekly, 2020. 2(5): p. 79–80.

4. Organization, W.H. There is a current outbreak of Coronavirus (COVID-19) disease. 2020; Available from: https://www.who.int/health-topics/coronavirus#tab=tab_1.

5. Zhang Rong, Y.Z., Wang Yuyan, Teng Zheng, Xu Wei, Song Wuhui, Cai Xia, Sun Zhiping, Gu Chenjian, Zhou Yanqiu, Chen Hongyou, Ye Rong, Han Wendong, Zhu Yunkai, Feng Fanghao, Li Chongshan, Zhang Xi, Qu Di, Fu Chen, Xie Youhua, Yuan Zhenghong, Isolation of a 2019 novel coronavirus strain from a coronavirus disease 19 patient in Shanghai. Journa of Microbes and Infections, 2020. 15(1): p. 15–20.

